# Urgent Transcatheter Edge-to-Edge Repair for Severe Mitral Regurgitation with Flail Leaflet in Critically Ill Patients

**DOI:** 10.1101/2023.03.25.23287602

**Authors:** Nimrod Perel, Itshak Amsalem, Or Gilad, Rafael Hitter, Tomer Maller, Elad Asher, Emanuel Harari, David Marmor, Shemy Carasso, Danny Dvir, Michael Glikson, Mony Shuvy

## Abstract

**Introduction:** Degenerative mitral valve disease (DMR) is a common valvular disorder, and flail leaflet due to ruptured chordae represents an extreme variation of this pathology. Ruptured chordae can present as acute heart failure which requires urgent intervention. Mitral valve surgery is the preferred mode of intervention; however, some patients represent as a significant surgical risk and are often deemed to be inoperable. We aim to characterize patients with ruptured chordae undergoing urgent transcatheter edge-to-edge repair (TEER), and to analyze their clinical and echocardiographic outcomes.

**Methods:** At a referral center for several hospitals across Israel, we screened all patients who underwent TEER. We included patients with DMR with flail leaflet due to ruptured chordae, and categorized them into elective and critically ill groups. We evaluated the echocardiographic, hemodynamic and clinical outcomes of these patients.

**Results:** The cohort included 49 patients with DMR due to ruptured chordae and flail leaflet, who underwent TEER. Seventeen patients (35%) underwent urgent intervention and 32 patients (65%) underwent an elective procedure. In the urgent group, the average age of the patient was 80.3, with 41.8% being female. Fourteen (82%) received noninvasive ventilation, while three patients (18%) required invasive mechanical ventilation. One patient died due to tamponade, while the other 16 patients all had successful reduction of ≥2 in the MR grade. Left atrial V wave decreased from 41.6 mmHg to 17.9 mmHg (p<0.001), and pulmonic vein flow pattern changed from reversal (68.8%) to a systolic dominant flow in all patients (p=0.001). After the procedure, 78.5% of patients were in New York Heart Association (NYHA) class I or II (p<0.001). There was no significant difference in the overall mortality between the urgent and elective groups, with similar survival rates of 6 months for each group.

**Conclusion:** TEER can be safe and feasible with favorable hemodynamic, echocardiographic and clinical outcomes in patients undergoing urgent intervention for flail mitral valve disease.

**Clinical Perspectives:**

- Mitral transcatheter edge-to-edge repair (TEER) is a proven therapy for patients with chronic mitral regurgitation, however data on patients with acute mitral regurgitation limited. This article focuses on acute TEER intervention and provides further information on the safety, feasibility and effectiveness of TEER in critically ill patients due to flail leaflet.
- The clinical finding supports the role of acute TEER intervention in acutely ill patients with acute mitral regurgitation due to ruptured chordae and flail leaflet.

## Introduction

Primary mitral regurgitation is a common valvular disease in which a primary lesion in any component of the valve apparatus can lead to mitral regurgitation (MR).^1^

MR is the second most common valvular disorder, with primary MR being almost twice as common as secondary MR. Degenerative mitral regurgitation (DMR) due to myxomatous degeneration has two distinct etiologies: fibroelastic deficiency with a focal lesion, or Barlow disease with diffuse involvement of both leaflets. The average age of patients with DMR is over 65; patients with fibroelastic deficiency tend to be older with a more acute presentation, while patients with Barlow disease are younger with a more chronic disease course.^2,3^

According to the Carpentier classification, DMR is categorized as Carpentier type 2 with leaflet or chordal degeneration leading to an excessive leaflet motion. Flail leaflet represents an extreme variation of this pathology, with excessive leaflet motion most frequently being caused by ruptured chordae. According to the American Society of Echocardiography (ASE) guidelines on non-invasive evaluation of native valvular regurgitation, flail leaflet is one of the specific criteria for severe MR and its presence often denotes severe MR.^4,5^

Patients with flail leaflet can present either acutely, with pulmonary edema and cardiogenic shock, or chronically with signs and symptoms of chronic heart failure. Despite surgery being the preferred mode of intervention, many of these patients are at significant surgical risk and are often deemed to be inoperable. Furthermore, in patients that undergo urgent surgical intervention, morbidity and mortality are significantly higher than in elective cases.^6^ As a result, many patients rely solely on medical treatment without having intervention, necessitating the need for a less invasive therapeutic strategy.

Transcatheter edge-to-edge repair (TEER) has been proven to be a safe alternative option for patients with chronic primary MR.^7^ Data on the use of urgent TEER for acutely ill patients in cardiogenic shock with significant MR, is scarce and originates from smaller cohorts, although the results are encouraging and show improved survival rates for patients undergoing TEER in these situations. However, most of the data on acute TEER intervention is based on functional MR, especially in the setting of acute myocardial infarction.^8,9^ Data on the use of acute TEER for patients with degenerative MR due to acute ruptured chordae and flail leaflet is limited. We aim to focus on patients that presented with acute symptoms and who underwent an urgent TEER procedure. In addition, in order to obtain a better understanding of this group, we compared their echocardiographic and clinical outcomes to individuals undergoing elective procedures.

## Methods

### Study population

We screened all patients who underwent TEER from January 2020 until October 2022 and followed them until January 2023. All patients underwent the procedure at a single medical center that serves as a referral center for several hospitals across Israel and which averages over 100 TEER procedures annually. Procedures are performed for both functional and degenerative MR, including patients who require urgent interventions.

Patients typically receive elective TEER procedures unless they develop HF symptoms that persist and necessitate urgent TEER intervention. In the current study, we focused on patients with severe MR with flail leaflet due to ruptured chordae and categorized them into elective and critically ill groups. Patients in the elective group presented with signs and symptoms of chronic heart failure, without acute decompensation that required urgent intervention. Patients in the urgent group were hospitalized for acute heart failure requiring an urgent procedure during that hospitalization. Additional patient stratification was done based on severity, ranging from non-invasive ventilatory assistance to cardiogenic shock with mechanical circulatory support. All patients were at high surgical risk, and after discussion were approved for TEER by the heart team.

### Echocardiographic assessment

Transthoracic echocardiography (TTE) was initially performed in all patients to assess the severity of the MR. Subsequently, all patients underwent transesophageal echocardiography (TEE) to further evaluate the mechanism of the regurgitation and to assess their suitability for TEER. Evaluation of severe MR was based on current ASE guidelines and was graded from 1-4 as mild, moderate, moderate-severe, or severe. Patients underwent transthoracic echocardiographic examination the day following the procedure; the severity of the MR was recorded using the same parameters as before the procedure, with the addition of measuring the post-procedural mitral gradient.

### Intervention

Transcatheter edge-to-edge repair was performed by the implantation of a “MitraClip®” device (Abbott, Abbott Park, IL, USA). The device system was delivered to the left atrium (LA) via transeptal puncture. The procedure was done with fluoroscopic and TEE guidance. TEE was used for grading the severity of the MR and for decision-making during the procedure (the implantation of an additional clip or the ending of the procedure). At least one clip was implanted in all patients. Additional clips were implanted to achieve a more significant MR reduction when deemed necessary by the operators. Left atrial V wave was measured and recorded before and after TEER implantation. Measurement after implantation was done while the TEE probe was still in place and before the iatrogenic shunt was unblocked (the removal of the clip delivery system) in order to provide more reliable measurements of LA pressure. We calculated the absolute reduction in the Left atrial V wave before and after the procedure. An assessment of the pulmonic systolic vein flow pattern was checked at the beginning of the procedure and after final clip implantation.

### Outcomes

We evaluated clinical outcomes, including the New York Heart Association (NYHA) functional class, before the procedure and at the first post-procedure follow up. We monitored the mortality rates at 30-days and 6-months post-procedure.

Echocardiographic outcomes included: post-procedure mitral gradient, pulmonic vein flow systolic pattern (the appearance of a systolic dominance pattern after the procedure), grade of MR reduction, and changes of the tricuspid insufficiency gradient.

### Statistical analysis

We evaluated all patients in the cohort for their baseline characteristics and their echocardiographic and hemodynamic parameters. We compared the parameters between the urgent and elective groups before and after the procedure. In addition, we compared the parameters before and after the procedure within the urgent group.

Logical checks were performed on the data for quality control purposes. Normally distributed continuous variables are presented as means (±standard deviations) and non-normally distributed continuous variables are presented as medians (interquartile ranges).

Comparisons of independent (e.g. elective vs. urgent procedures) continuous variables were made using independent sample T-tests. Comparisons of continuous variables before and after the procedures were carried out by using paired samples T-tests. Categorical variables were compared using the Chi-Square test. When necessary, due to non-normal distributions, non-parametric tests were used.

All analyses were conducted using SPSS statistical software (version 26.0) and Rstudio (version 2022.07.2+576). All statistical tests were two-sided, and significance was determined at a p-value of 0.05.

## Results

From a cohort of 236 patients who underwent TEER implantation between January 2020 and October 2022, we identified 49 patients who had DMR with ruptured chordae and flail leaflet. Thirty-two patients out of 49 (65%) underwent an elective procedure, while the other 17 patients (35%) underwent urgent intervention with TEER as a non-elective procedure during their hospitalization due to heart failure (Figure 1). Follow up time ranged between 3-36 months with a mean of 17.16 (±8.95) months.

**Figure 1:** Trial Inclusion Criteria SZMC -TMVR - Shaare Zedek medical center transcatheter mitral valve repair; FMR - functional mitral regurgitation; DMR - degenerative mitral regurgitation; OMT - optimal medical therapy; TEER - transcatheter edge to edge repair

The average age of the cohort was 79.7 ± 9 years, with 17 patients being female (34.6%). Hypertension was the most common comorbidity, with 40 patients being affected by this (83.3%). This was followed by dyslipidemia and diabetes mellitus which were present in 29 and 11 patients respectively (60.4% and 22.9%). There was no significant difference in most of the baseline characteristics between the patients in the elective and urgent groups. There was a significant difference in the NYHA class between the two groups. All 17 patients in the urgent group were NYHA IV compared with three out of 32 patients (9.6% p<0. 001) in the elective group (Table 1).

**Table 1.**
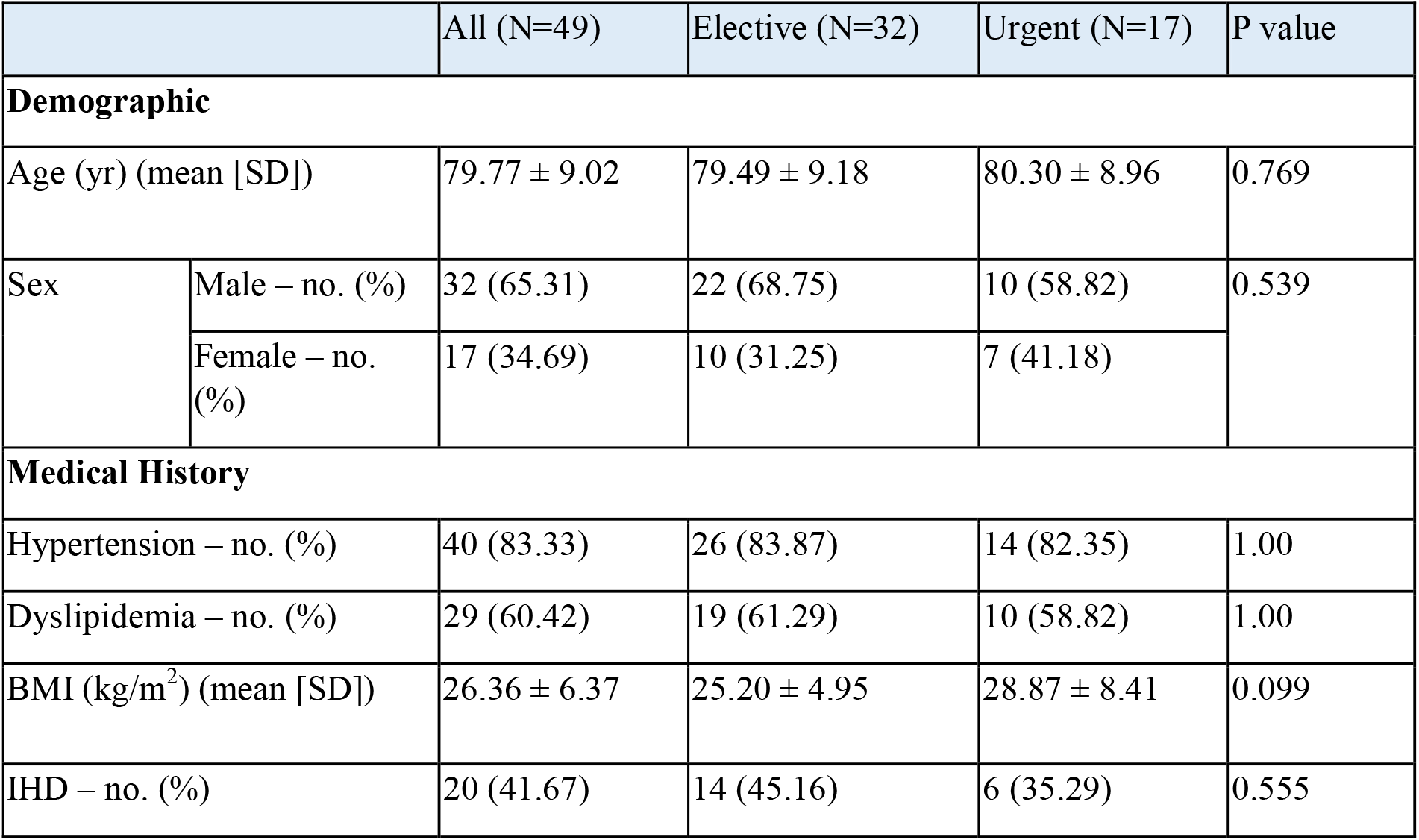
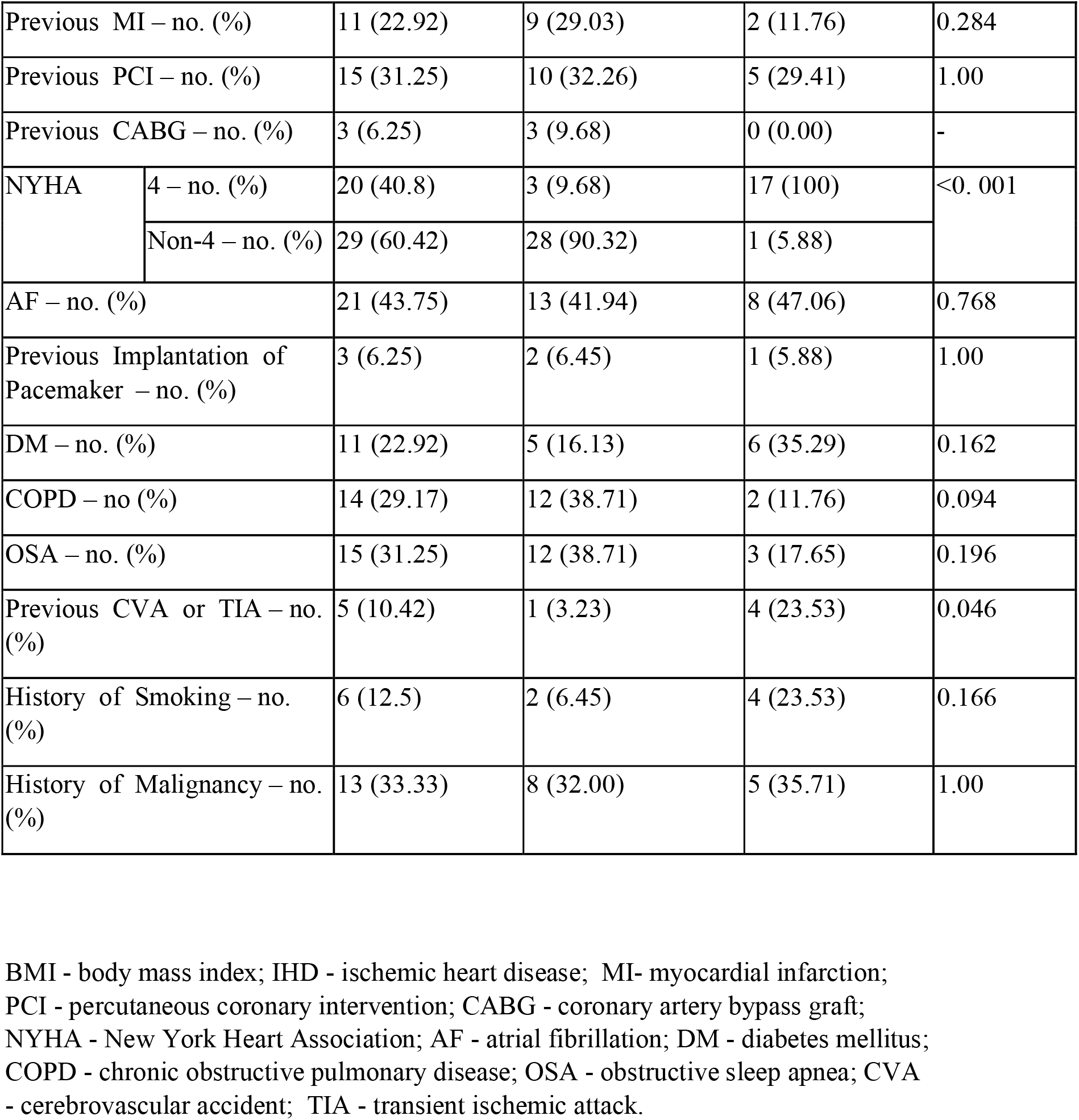
Baseline Characteristics

|  |  | All (N=49) | Elective (N=32) | Urgent (N=17) | P value |
| --- | --- | --- | --- | --- | --- |
| Demographic |  |  |  |  |  |
| Age (yr) (mean [SD]) |  | 79.77 ± 9.02 | 79.49 ± 9.18 | 80.30 ± 8.96 | 0.769 |
| Sex | Male – no. (%) | 32 (65.31) | 22 (68.75) | 10 (58.82) | 0.539 |
|  | Female – no. (%) | 17 (34.69) | 10 (31.25) | 7 (41.18) |  |
| Medical History |  |  |  |  |  |
| Hypertension – no. (%) |  | 40 (83.33) | 26 (83.87) | 14 (82.35) | 1.00 |
| Dyslipidemia – no. (%) |  | 29 (60.42) | 19 (61.29) | 10 (58.82) | 1.00 |
| BMI (kg/m<sup>2</sup>) (mean [SD]) |  | 26.36 ± 6.37 | 25.20 ± 4.95 | 28.87 ± 8.41 | 0.099 |
| IHD – no. (%) |  | 20 (41.67) | 14 (45.16) | 6 (35.29) | 0.555 |
| Previous MI – no. (%) |  | 11 (22.92) | 9 (29.03) | 2 (11.76) | 0.284 |
| Previous PCI – no. (%) |  | 15 (31.25) | 10 (32.26) | 5 (29.41) | 1.00 |
| Previous CABG – no. (%) |  | 3 (6.25) | 3 (9.68) | 0 (0.00) | - |
| NYHA | 4 – no. (%) | 20 (40.8) | 3 (9.68) | 17 (100) | <0.001 |
|  | Non-4 – no. (%) | 29 (60.42) | 28 (90.32) | 1 (5.88) |  |
| AF – no. (%) |  | 21 (43.75) | 13 (41.94) | 8 (47.06) | 0.768 |
| Previous Implantation of Pacemaker – no. (%) |  | 3 (6.25) | 2 (6.45) | 1 (5.88) | 1.00 |
| DM – no. (%) |  | 11 (22.92) | 5 (16.13) | 6 (35.29) | 0.162 |
| COPD – no (%) |  | 14 (29.17) | 12 (38.71) | 2 (11.76) | 0.094 |
| OSA – no. (%) |  | 15 (31.25) | 12 (38.71) | 3 (17.65) | 0.196 |
| Previous CVA or TIA – no. (%) |  | 5 (10.42) | 1 (3.23) | 4 (23.53) | 0.046 |
| History of Smoking – no. (%) |  | 6 (12.5) | 2 (6.45) | 4 (23.53) | 0.166 |
| History of Malignancy – no. (%) |  | 13 (33.33) | 8 (32.00) | 5 (35.71) | 1.00 |

BMI - body mass index; IHD - ischemic heart disease; MI- myocardial infarction; PCI - percutaneous coronary intervention; CABG - coronary artery bypass graft; NYHA - New York Heart Association; AF - atrial fibrillation; DM - diabetes mellitus; COPD - chronic obstructive pulmonary disease; OSA - obstructive sleep apnea; CVA - cerebrovascular accident; TIA - transient ischemic attack.

All patients in the urgent group needed ventilatory assistance. Thirteen patients (76.4%) were treated by non-invasive ventilatory support, while four patients (23%) needed invasive mechanical ventilation. Two patients (11%) were in severe cardiogenic shock and required pharmacological and mechanical assistance. Cardiogenic shock was defined as systolic blood pressure <90 mmHg with signs of end organ hypoperfusion.^10^ The baseline echocardiographic parameters of the group included the presence of a flail in the posterior leaflet in 12 patients (70.5%), and a bi-leaflet flail in one patient. Thirteen patients (76.4%) had an A2-P2 pathology, and four patients (24.6%) had a non-A2-P2 pathology. The average flail gap was 7.1 mm, with a range between 0.3 mm and 1.1 mm.

Postprocedural hemodynamic and echocardiographic parameters showed a significant improvement in the MR grade. All patients in the urgent group had a grade of +4 MR before the procedure, with a reduction to an average grade of +1.3 MR (Figure 2). In the paired analysis, the average left atrial V wave before the procedure was 41.6 mmHg, with a statistically significant change to an average left atrial V wave of 17.9 mmHg after the procedure (p<0.001) (Figure 3). The pulmonic vein flow patterns were systolic flow reversal in 11 patients (68.8%) and systolic blunting in four patients (25%). The pattern of flow changed to a systolic dominance pattern in all patients after the procedure (p=0.001) (Figure 4). The systolic pulmonary arterial pressure average was 52.6 mmHg before the procedure and was decreased to an average value of 44.25 mmHg (p=0.06) (Figure 5). The improvement of the hemodynamic and echocardiographic parameters were similar in the elective group (Tables 2-3). The average number of clips implanted was similar in both groups (1.69 in the elective group vs 1.88 in the urgent group p=0.28). In the urgent group, the type of clips used was XTW (30.0%), XT (26.6%), NTW (23.4%), NT (20.0%)

**Figure 2:** Transthoracic echocardiography evaluation of the MR grade before the procedure and at 1 day after the procedure in the urgent group MR- Mitral Regurgitation; TEER - transcatheter edge to edge repair

**Figure 3:** Left atrial V wave change before the procedure and immediately after the clip implantation in the urgent group

**Figure 4:** Transesophageal echocardiography evaluation of pulmonic vein flow pattern before the procedure and after the clip implantation in the urgent group TEER - transcatheter edge to edge repair

**Figure 5:** Transthoracic echocardiography evaluation of systolic pulmonary artery pressure before the procedure and at 1 day after the procedure in the urgent group TEER - transcatheter edge to edge repair

**Table 2.**
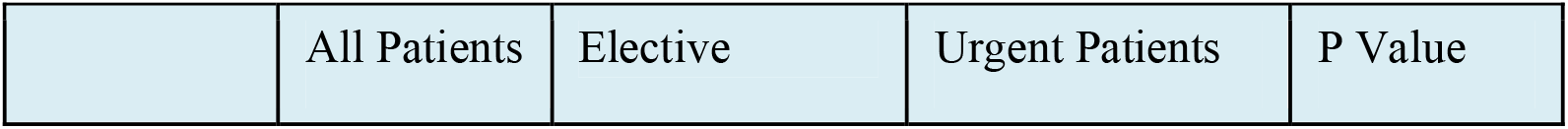
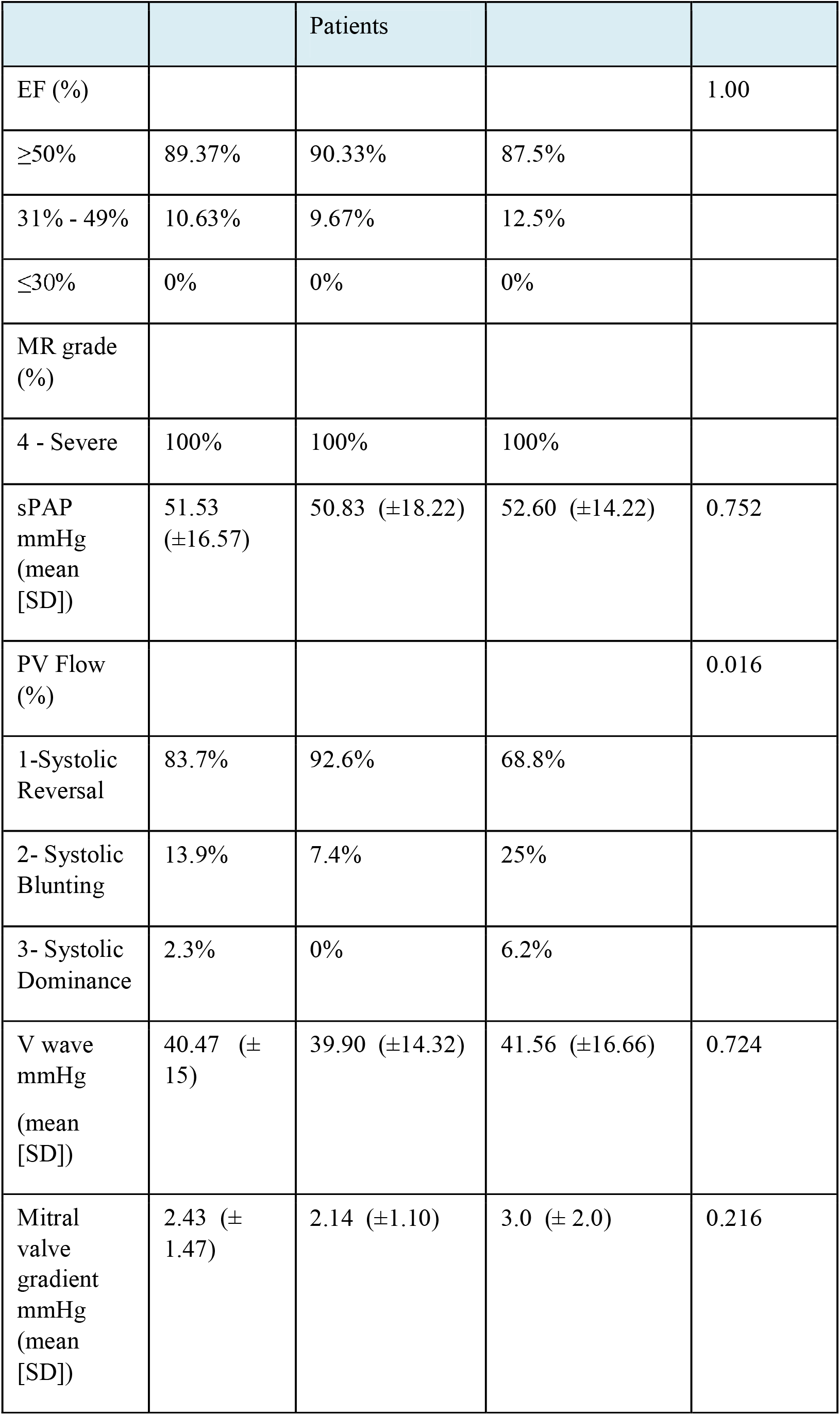
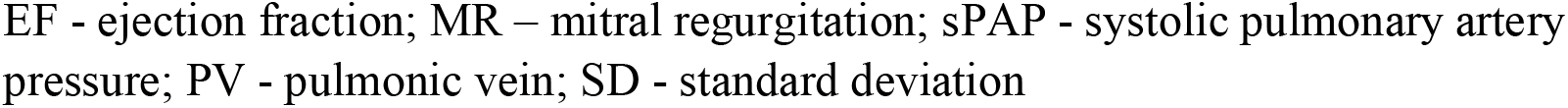
Echocardiographic and Hemodynamic Parameters Before TEER in All Groups

**Table 3.** Echocardiographic and Hemodynamic Parameters After TEER in All Groups

|  | All Patients | Elective Patients | Urgent Patients | P Value |
| --- | --- | --- | --- | --- |
| MR grade (%) |  |  |  | 0.498 |
| 0 -Trace | 4.3% | 3.2% | 6.2% |  |
| 1- Mild | 40.4% | 32.3% | 56.2% |  |
| 2 - Moderate | 44.7% | 51.6% | 31.2% |  |
| 3 - Moderate Severe | 8.5% | 9.7% | 6.2% |  |
| 4 - Severe | 2.1% | 3.2% | 0% |  |
| sPAP mmHg (mean [SD]) | 43.13 (±14.43) | 42.53 (±14.62) | 44.25 (±14.46) | 0.705 |
| Delta sPAP mmHg (mean [SD]) | -7.44 (± 15.97) | -6.82 (± 16.69) | -8.43 (± 15.33) | 0.773 |
| PV flow (%) |  |  |  | 0.2 |
| 1 - Systolic Reversal | 2.3% | 3.6% | 0% |  |
| 2 - Systolic Blunting | 9.1% | 14.3% | 0% |  |
| 3 - Systolic Dominance | 88.6% | 82.1% | 100% |  |
| Left atrial V wave mmHg (mean [SD]) | 19.9 (±7.08) | 17.73 (±7.31) | 17.93 (±6.84) | 0.927 |
| Delta Left atrial V wave mmHg (median [IQR]) | -20 [17.0] | -20 [16.0] | -20 [19.0] | 0.907 |
| Mitral Valve Gradient mmHg (mean [SD]) | 5.14(±2.73) | 5.29 (±3.27) | 4.86 (±1.21) | 0.743 |
| Delta Mitral Valve Gradient mmHg (mean [SD]) | 2.71 (±3.00) | 3.14 (±3.48) | 1.86 (±1.57) | 0.368 |
| Number of Clips Implanted Number of clips (mean (SD)) | 1.75 (0.56) | 1.69 (0.53) | 1.88 (0.62) | 0.283 |

EF - ejection fraction; MR - mitral regurgitation; sPAP - systolic pulmonary artery pressure; PV - pulmonic vein; SD - standard deviation

Clinically, in the urgent group there was a significant improvement in the NYHA class (Figure 6). Seventy-eight percent of the patients improved to NYHA I or II at the first follow-up visit (p <0.001). Sixteen patients in the urgent group (94.1%) were alive at least six months following the procedure, whilst one patient died during the procedure due to cardiac tamponade that occurred during the transeptal puncture (Table 4). There was no significant difference in the improvement of the NYHA class when compared with the elective group.

**Figure 6:** NYHA class before the procedure and after the procedure at a follow up evaluation in the urgent group NYHA – New York Heart Association

**Table 4.**
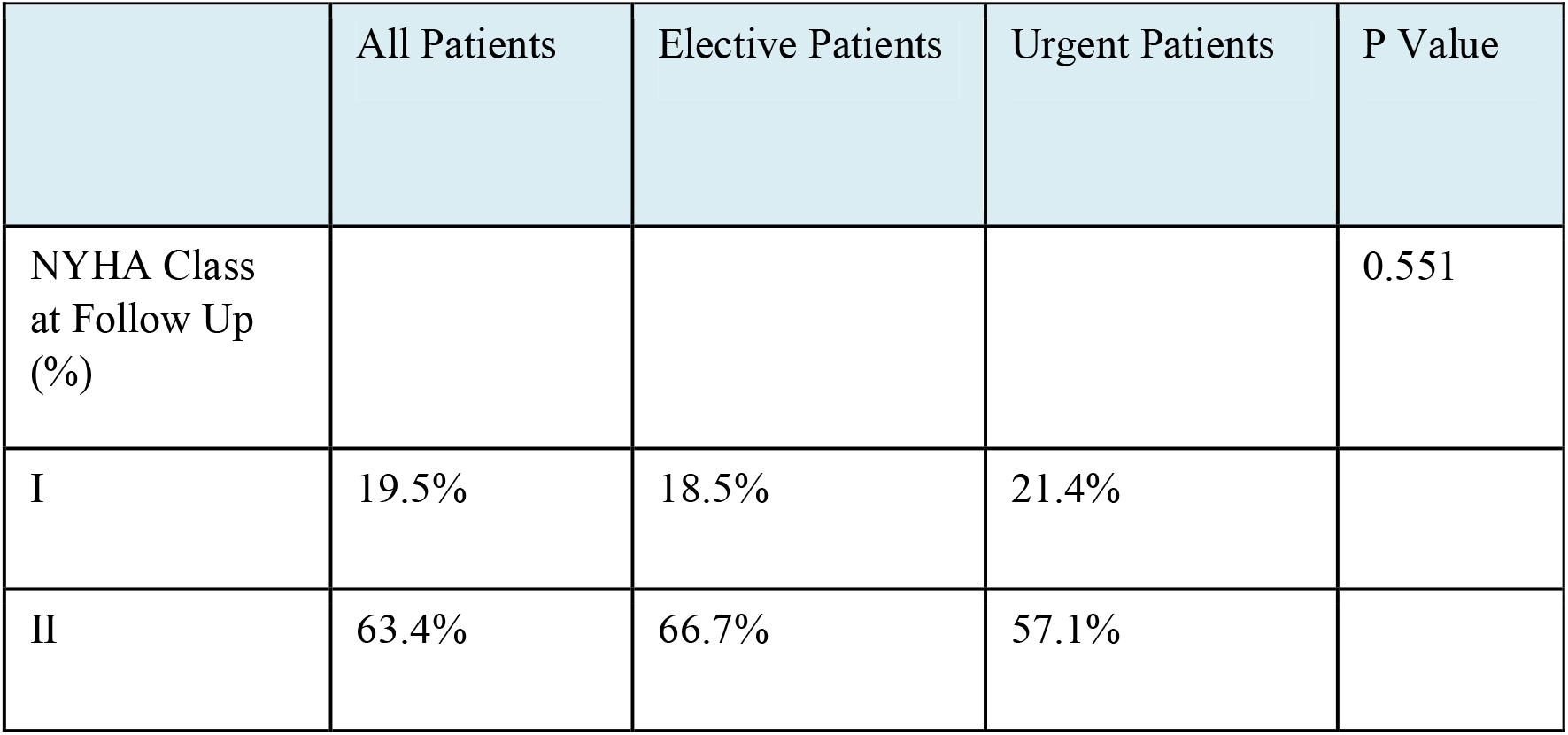
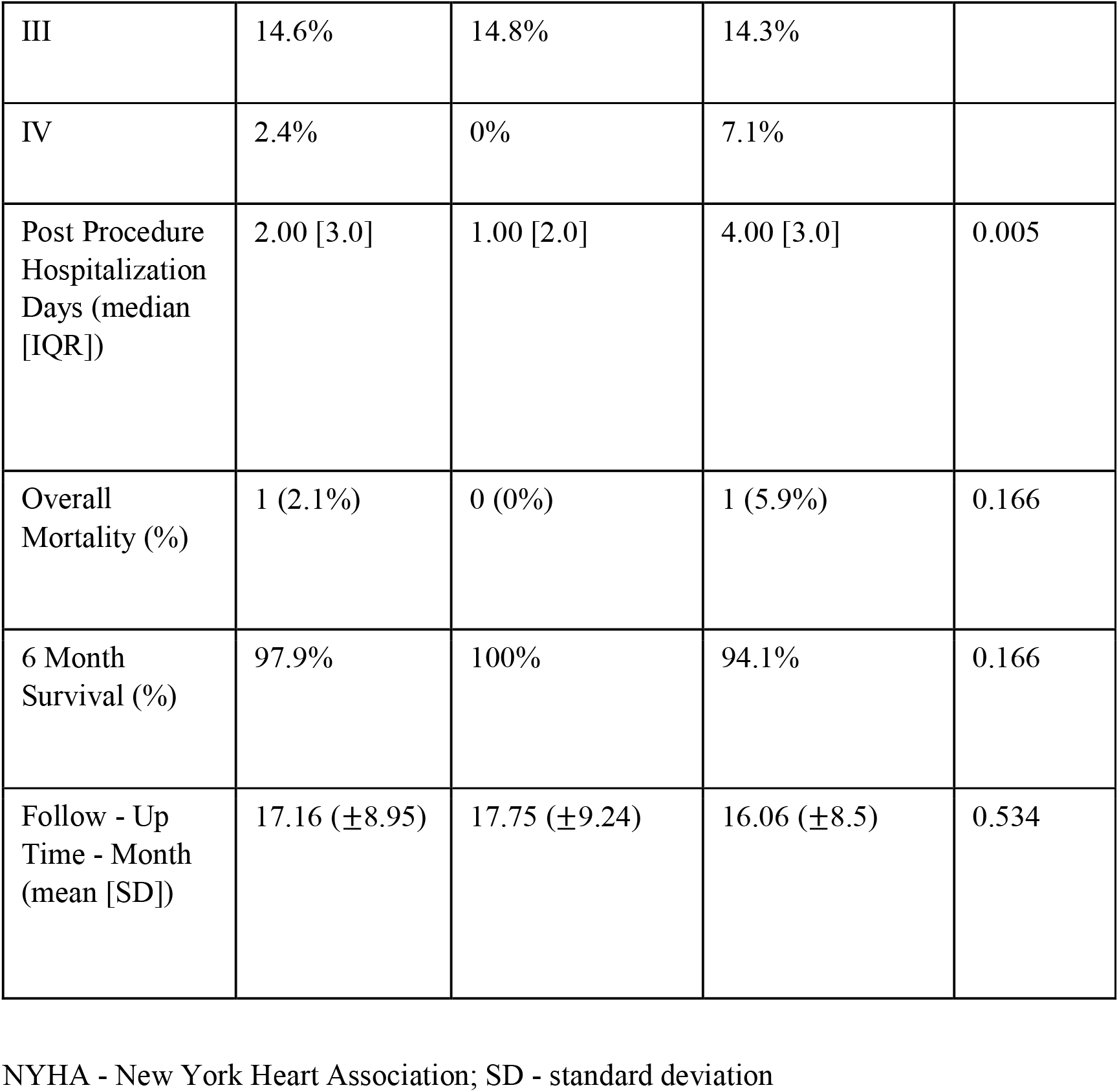
Clinical Outcome of Patients in All Groups

|  | All Patients | Elective Patients | Urgent Patients | P Value |
| --- | --- | --- | --- | --- |
| NYHA Class at Follow Up (%) |  |  |  | 0.551 |
| I | 19.5% | 18.5% | 21.4% |  |
| II | 63.4% | 66.7% | 57.1% |  |
| III | 14.6% | 14.8% | 14.3% |  |
| IV | 2.4% | 0% | 7.1% |  |
| Post Procedure Hospitalization Days (median [IQR]) | 2.00 [3.0] | 1.00 [2.0] | 4.00 [3.0] | 0.005 |
| Overall Mortality (%) | 1 (2.1%) | 0 (0%) | 1 (5.9%) | 0.166 |
| 6 Month Survival (%) | 97.9% | 100% | 94.1% | 0.166 |
| Follow - Up Time - Month (mean [SD]) | 17.16 ($\pm 8.95$) | 17.75 ($\pm 9.24$) | 16.06 ($\pm 8.5$) | 0.534 |

NYHA - New York Heart Association; SD - standard deviation

## Discussion

The natural history of MR due to flail leaflet carries a high risk of morbidity and mortality as a result of left ventricular dysfunction, heart failure and death. The clinical outcome of patients with flail leaflet treated medically without intervention has been significantly worse than those who underwent surgical correction.^11,12^

The recommended mode of correction is a surgical repair of the valve while percutaneous edge-to-edge repair can be a safe and effective alternative in high-risk patients. The current recommendation refers mainly to patients with chronic MR, while the recommendation for patients with acute MR is based on pathological conditions such as papillary muscle rupture or infective endocarditis in which prompt restoration of the integrity of the valve is essential for survival.^1,7^ However, patients with acute MR are a very high surgical risk, and many cannot undergo surgery. According to Lorusso *et al* who evaluated the outcomes of 279 patients who underwent emergency surgery for: severe acute MR brought on by an acute myocardial infarction, degenerative mitral valve disease, or acute endocarditis, the 30-day mortality rate was 22.5% overall, and 14.8% among patients with degenerative valve disease. The high mortality rates in patients with degenerative mitral valve who often have preserved left ventricle systolic function without severe comorbidities, emphasizes the non-negligible risk of surgery in the acute setting and raises the need for a lower risk intervention, even for patients with primary MR.^6^

In this study we demonstrate and characterize the echocardiography, hemodynamic and clinical outcomes of 49 elderly patients (average age was 80 years old) with DMR due to flail leaflet, that underwent TEER. We further divided the cohort of patients into acute patients requiring an urgent TEER procedure, and patients with chronic symptomatic MR who underwent elective procedures. We made a comparison between the elective and urgent groups, aiming to compare the outcome between the groups. We then focused on the urgent group, describing in more detail their clinical and echocardiographic parameters. In this group, we compared the parameters before and after the procedure, aiming to show and emphasize the high rate of procedural success and ultimately their positive clinical outcomes. We showed the procedure to be safe, achieving a high rate of procedural success in a high-risk elderly patient population.

Our clinical and echocardiography outcomes are similar to other retrospective studies involving patients with chronic DMR who were at a high surgical risk and who underwent TEER.^13,14^ However, in our study, we exclusively described patients with flail leaflet rather than any other degenerative pathology. The flail mechanism usually denotes a severe MR and often implies a more complicated procedure (e.g. large coaptation gap, etc.).^5^ The use of newer devices such as the fourth generation Mitraclip or Pascal ACE and the ability to perform independent leaflet grasping allowed the treatment for more challenging mitral valve anatomies including wide coaptation gaps, large redundant tissue and large flail gap.

In contrast to chronic MR, in which there is time for a compensating mechanism and hence a more gradual deterioration, in acute MR patients usually present with severe respiratory distress, often with pulmonary edema and even cardiogenic shock.^15^

There is currently less data on TEER in patients with acute MR, although emerging data shows favorable results in patients with functional MR (mostly secondary to ischemic insult).^16–18^ However, information describing patients with acute primary MR due to ruptured chordae, who undergo an acute TEER procedure, is limited.

The average age and comorbidities of our cohort were similar to previously mentioned DMR patients who underwent TEER. However, their acute presentation and severe current illness put them at a higher risk for surgery and mortality in general. We demonstrate a high rate of MR reduction, with most patients having >2 grade reduction. This reduction in the severity of MR addresses the main concern regarding percutaneous repair versus surgical repair, with surgical repair achieving a greater MR reduction.^19^ Furthermore, we showed a significant clinical benefit by significantly improving the patient’s NYHA class. This clinical improvement supports the previous finding that procedural success with significant MR reduction is the major determinant of clinical outcome.^20,21^

The mortality rate of patients undergoing urgent TEER was non-inferior to the mortality rate of the elective patients with chronic MR. These findings further support the safety and clinical benefit of TEER in those patients who cannot undergo surgery.

### Limitation

This analysis is based on retrospective observational data without a control group and is limited in size. Because the study’s cohort is based on patients undergoing TEER at a high-volume single center with an experienced team, the results of the procedure and the outcomes of the patients cannot be generalized to all other centers performing TEER. In addition, since several patients were referred from other hospitals, some of their data could not be retrieved and is therefore missing from the overall data of the study. Our findings support the role of TEER in these acutely ill elderly patients who do not have other therapeutic options. Nevertheless, for patients undergoing TEER with suboptimal results, either as an urgent or elective procedure, a secondary procedure with surgery usually mandates a replacement of the valve, rather than a repair.

## Conclusion

TEER can be safe and effective in patients with DMR, including patients presenting with acute severe MR. In acutely ill patients with prohibitive surgical risk, TEER can serve as a good therapeutic alternative, providing a bridge to recovery.

## Data Availability

all data avilable in this manuscript is original

